# Effect of HPV Vaccine Introduction at 9-10 years: A Randomized Clinical Trial

**DOI:** 10.64898/2026.09.24.26363898

**Authors:** Allison Kempe, Sitaram Vangala, Christina Albertin, Sean T. O’Leary, Brenda Beaty, Alison W. Saville, Dennis Gurfinkel, Sharon G. Humiston, Shen Nagel, Jasjit Singh, Charles Golden, Heide Woo, Emma J. Clark, Peter G Szilagyi

## Abstract

**Importance:** In the US, Human papillomavirus (HPV) related cancers cause significant morbidity and mortality, yet HPV vaccine is underutilized. Vaccination is recommended routinely at age 11-12 years but can be given at 9 years. There is controversy about whether earlier introduction would increase completion of the HPV series before by age 13 years or whether it might decrease rates of delivery of other routinely recommended vaccines at 11-12-years [quadrivalent meningococcal conjugate (MenACWY) vaccine, and tetanus-diptheria-acellular pertussis (Tdap) vaccine].

**Objective:** To determine whether introduction of the HPV vaccine at ages 9-10 versus 11-12 years, results in earlier median age of series completion (primary outcome) and higher rates of initiation and completion by age 13 years. We also examined the effect of earlier introduction of HPV on rates of Tdap and MenACWY initiation and completion by age 13 years.

**Design:** Two-arm cluster-randomized trial involving 31 pediatric practices in Colorado and California between 10/2021-4/2026. Practices were randomized using covariate constrained randomization within each state to Intervention (introduction of HPV vaccination at ages 9-10) or Control (introduction at ages 11-12). Both study groups received similar annual training about best practices for HPV vaccination with recommended initiation age as the only difference.

**Results:** Age at HPV series completion was substantially earlier in the Intervention group (median 12.2 v. 13.3 years; adjusted hazard ratio [aHR] 2.37; 95% CI 1.94-2.89). Key secondary outcomes were also improved in the intervention group: age at series initiation (median 11.0 v. 11.8 years; aHR 2.19; 95% CI 1.84-2.62), series completion by 13 years (61.6% v. 45.1%; RR 1.37; 95% CI 1.04-1.74), and vaccine initiation by age 13 years (77.1% v. 67.0%; RR 1.15; 95% CI 1.00-1.33). The intervention group was more likely to receive Tdap (aHR 1.47;95%CI 1.13, 1.92) during the trial; receipt of both vaccines by age 13 years did not differ.

**Conclusions:** Earlier introduction of HPV vaccination resulted in earlier age of series completion, and more adolescents initiating and completing the series by age 13 without decreasing rates of other routinely recommended vaccines. Our findings support a preferential recommendation at 9-10 years.

**Trial registration number:** NCT04722822 ClinicalTrial.gov

**Key Points:** In the US human papillomavirus (HPV) vaccination is routinely recommended at age 11-12 but can be given at 9-10 years. This RCT compared practices randomized to initiation at age 9-10 vs 11-12 years. The 9-10 group vs the 11-12 group completed the HPV series over a year earlier and had their first HPV dose 8 months earlier. By age 13 years 77% vs 67% had initiated vaccination and 61.6% vs 45.1% had completed the series. Other adolescent vaccines were received at the same or higher rates in the 9-10 group. Preferential recommendation for HPV at 9-10 years should be considered.

## Introduction

Human papillomavirus (HPV) infection and subsequent HPV-related cancers remain a major public health problem across the US and globally. Nearly all cervical cancers, 91% of anal, 63-75% of genital cancers, and 70% of oropharyngeal cancers are due to HPV infection. In the US, 37,300 new HPV-associated cancers occur annually, with oropharyngeal and cervical cancers most common.^1,2^

Highly effective and safe vaccines against HPV infection have been licensed in the US since 2006. The 9-valent HPV vaccine is the only vaccine marketed in the US since 2016, with protection against 90% of HPV-attributable cancers.^3^ Clinical trials showed high effectiveness in preventing precancerous cervical lesions.^4-6^ Large population-based studies show 86-88% lower risks of cervical cancer if vaccinated during adolescence, with early evidence of effectiveness for oropharyngeal^7^ cancers and for penile and anal cancer^8^ among men. Importantly, vaccine effectiveness has been shown to be long-lived, especially when given at an early age, with antibody persistence of approximately 98% at 10 years among those vaccinated aged 9-14 years.^9^ Vaccine safety is supported by prelicensure vaccine trials and by 15+ years of post-licensure monitoring, with over 135 million doses of HPV vaccine distributed in the US through 2021.^2^ Because the vaccine should be given prior to sexual activity, the National Committee for Quality Assurance’s Healthcare Effectiveness Data and Information Set (HEDIS) measure for adolescent vaccination, used by >90% of US health plans as a performance measure, includes HPV series completion by age 13 years.^10^

At the time this trial began in 2020, the Advisory Committee on Immunization Practices (ACIP), which advises the Centers for Disease Control and Prevention (CDC), recommended HPV vaccination to begin routinely at age 11-12 but stated, “Vaccination can be given starting at age 9 years.”^11^ Two doses were recommended to complete the series if vaccination was begun at <15 years of age, with three doses for initiation after 15 years. Despite the effectiveness and safety of HPV vaccines, in 2024, only 78% of 13–17-year-olds had received one HPV vaccination and only 63% had completed the series per ACIP guidelines,^12^ far below coverage for other recommended adolescent vaccines.

Considerable research has focused on increasing HPV coverage by improving clinician communication, reminder-recall and practice-based quality improvement efforts.^13-25^ Although some interventions have shown substantial effectiveness, (e.g., 6.8% decreases in missed vaccination opportunity rates^15^ or increases of 9.5% in HPV initiation^17^) they generally are multicomponent interventions that can be complex for practices to implement. In recent years there has been increasing interest in shifting initiation of HPV vaccination to ages 9-10 yrs with potentially modest effort for practices. Retrospective studies and multi-component interventions that included initiation at 9-10 noted associations with higher completion^26^ and commentaries have advocated early initiation.^27,28^ However, no prior study involved a direct comparison of the effect of age of initiation without confounding with other concurrent interventions, and most clinicians still initiate at ages 11-12 years.^29,30^

Some have expressed concern that changing the age of HPV initiation would undermine the 11-12 year “adolescent platform”^31^ which was first recommended by ACIP and the AAP in 1996 to encourage routine health care and catch for recommended childhood vaccines.^32^ It was subsequently expanded as new vaccines were added to the routine schedule, including the quadrivalent meningococcal conjugate (MenACWY) vaccine,^33^ the tetanus-diptheria-acellular pertussis (Tdap) vaccine^34^ and the HPV vaccine.^35^ The bundling of vaccine administration at 11–12-years has been temporally associated with increases in adolescent vaccination rates and preventive care to young adolescents,^31,36^ although it is not clear if this is attributable to the platform.

The rationale for this trial was to provide high-grade evidence to guide recommendations for age of initiation of HPV vaccine. Our primary hypothesis was that age of series completion would be lower if vaccination was initiated at 9-10 years rather than 11-12 years. We also hypothesized that a practice policy change to introduce HPV vaccination at ages 9-10 would translate into higher series completion rates by age 13 and earlier average age of initiation of HPV vaccination (secondary outcomes}. Because of concern of the potential impact of earlier initiation on other adolescent vaccines, we also compared delivery of MenACWY and Tdap between study arms.

## Methods

This study received approval by the Colorado Multiple Institutions Review Board as an expedited review.

### Study design

This 2-arm cluster-randomized pragmatic comparative effectiveness trial was conducted in 31 urban primary care practices in Colorado (Denver Metro) and California (Los Angeles and Orange Counties) between October 2021 and April 2026. Practices were randomized within each state to Intervention (introduction of HPV vaccination at ages 9-10) or Control (continued introduction at ages 11-12).

### Study populations

In Colorado, 17 study practices were recruited from a Practice Based Research Network (COCONet)^37^; in California, 14 were recruited from two clinic systems (University of California Los Angeles Health System [UCLA Health] and Children’s Hospital Orange County [CHOC]).

These practices and health systems were selected because they incorporated different types of practices (mostly independent practices in CO, healthcare system-owned practices in CA), patient payer status and parts of the country. Eligibility criteria for practices included having ≥2 clinicians, an active roster of 100+ adolescents age 9-13, being a Pediatric or Med-Peds practice, and a routine policy of initiating HPV vaccine at ≥11 years (verified by practice managers). All clinicians were offered MOC Part IV credit to incentivize participation.

### Randomization

Practices were randomized separately (**Figure 1** CONSORT diagram) within state (and within health system in CA) using covariate constrained randomization to balance groups with respect to key variables associated with outcomes: number of adolescents 9-13 years by EHR (corresponded to practice size), baseline series completion rates of 13-year-olds (verified by EHR), and percentage of patients with Medicaid (self-report by practice managers).

**Figure 1:**
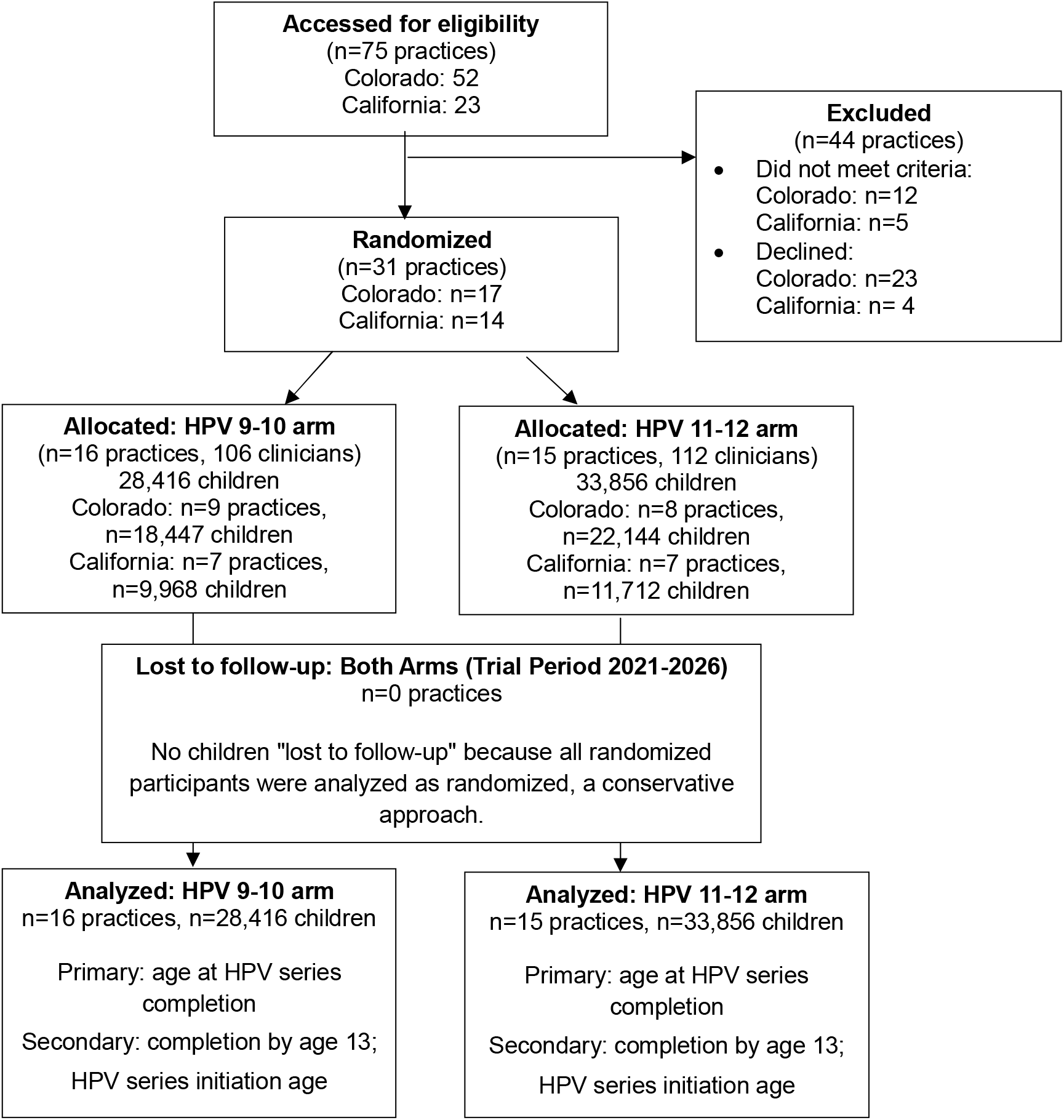
CONSORT Flow Diagram. CO = Colorado; CA = California. Randomization unit: practice (cluster). Unit of analysis: individual child (EHR data). ClinicalTrials.gov: NCT04722822. Funded by NCI R01CA240649.

### Interventions

The interventions were identical between study arms with respect to type of training, number of meetings with practices, time spent with the study team, and the type of feedback to practices. Practices in both arms received a single online training, repeated annually, about best practices for improving clinician communication and workflow enhancements for HPV vaccination. The training was adapted from previous trials studying interventions to improve HPV vaccination. Improved communication recommendations included brief training in presumptive recommendations^21^ and motivational interviewing when parents had concerns.

Workflow enhancement recommendations included vaccinating at all visits, provider and nurse prompts, standing orders, and posting the practice’s adolescent vaccine schedule in examination rooms. For the intervention arm, training included potential benefits of HPV vaccine initiation at 9-10 years and suggested wording about how to initiate this conversation including a statement that initiation of HPV vaccination at this age was now routine in the practice and supported by national recommendations, and that the change would reduce the number of vaccinations needed at age 11.

We did not require practices to make any of the recommended best practice recommendations and did not track their delivery practices but asked them to adhere to initiating the series at their randomly assigned age (9-10or 11-12 years). Therefore, the only intervention that differed between the control and intervention practices was the age of introduction. Each practice identified a study champion at the study onset who received a **$**400 yearly stipend for assisting with data collection, scheduling meetings and conducting surveys as part of MOC credit.

We deployed to both study arms: (a) online clinician surveys at 1, 6, 12, 18, 24, 30 and 36 months to understand feasibility, acceptability, parental hesitancy/interest, and discussion length and content; (b) feedback reports (EHR data) to practices about their captured HPV vaccination opportunities at 6, 12, 24 and 36 months (used for MOC); (c) 30-60 minute virtual meetings at baseline, 12, and 24 months to review the study and show feedback reports; and (d) wall posters if practices desired, displaying the adolescent vaccination schedule including HPV vaccine initiation at either 9-10 or 11-12 years. We tracked participation in training and annual repeat modules (>80% completion for both). No EHRs or state Immunization Information Systems changed their HPV vaccine alerts during the study.

### Data

In California, we extracted all data centrally from both health systems. In Colorado, 14 practices data pulled centrally via an integrated EHR; three practices provided practice-level data. Patient data were extracted from EHR data at the end of the study (November 2025 for Colorado and April 2026 for California). Extraction dates were based on each practice’s start date. Data variables collected included date of birth, sex, visits and visit dates, vaccines administered and dates, and insurance. Eligible patients included those (1) ages 9-10.5 years (i.e. those amenable to the intervention) at baseline who had an in-person clinic visit within 3 years prior to the study plus (2) patients with at least one visit during the study at age 9 to 10.5 years. We chose a 3-year lookback to account for potential reduction of in-person clinic visits during the COVID-19 pandemic. We followed these patients through time until the initial 9-year-olds reached age 13.0 years.

### Statistical analysis

We computed descriptive statistics for patient and practice characteristics, stratifying by treatment arm. We also assessed the visit-level rate at which vaccination opportunities (receipt of an HPV vaccine by an eligible child) were captured for 9–10-year-olds to assess fidelity. To test the primary study hypothesis that eligible patients in the 9-10 practices have earlier HPV series completion relative to those in 11-12 practices, we used a Cox proportional hazards regression model to compare age at vaccination between study arms. Patients not completing the series were administratively censored at the end of the study. The model adjusted for patient age, gender, state, and practice-level randomization balancing factors. Clustering of patients within practices was accounted for using robust covariance estimators.

To estimate the proportion completing the HPV series by age 13, estimates of the survival curve for age at HPV series completion were computed for each treatment arm using Kaplan-Meier curves for the primary outcome. Standard errors were estimated using the cluster bootstrap, with resampling at the practice level. Because recommendations might be changed to a single dose of HPV vaccine during adolescence, we repeated each analysis for initiation rather than completion. We evaluated, using interaction effects in regression models, whether patient or practice characteristics included in the regression analysis influenced intervention effectiveness. We replicated the same analyses for Tdap and MenACWY vaccines, assessing time-to-vaccination and receipt of each vaccine by age 13 years as previously described. A 0.05 significance level was used for hypothesis testing. A sensitivity analysis was performed limiting the sample to the children age 9-10 at baseline. Statistical analyses were performed using R version 4.5.1 and SAS version 9.4.

## Results

The intervention and control groups were similar with respect to practice-level variables included in the covariate constrained randomization (**Table 1**). Also shown are patient-level data, including age and gender. Race and ethnicity were not uniformly collected by participating practices and were not included. Captured HPV vaccination opportunity rates for 9–10-year-olds in the control group were 1-3% of eligible visits, in contrast to high and rising rates in the intervention group, demonstrating a high degree of fidelity (**Figure S1)**.

**Table 1.** Sample Characteristics of Practices and Participating Patients.

|  | Combined |  |  | Colorado |  |  | California |  |  |
| --- | --- | --- | --- | --- | --- | --- | --- | --- | --- |
| Mean (SD) | All | 9-10 | 11-12 | All | 9-10 | 11-12 | All | 9-10 | 11-12 |
| Baseline Practice Characteristics | (N=31) | (N=16) | (N=15) | (N=17) | (N=9) | (N=8) | (N=14) | (N=7) | (N=7) |
| Up-to-Date Rate (%) | 28.3<br>(17.6) | 28.8<br>(18.0) | 27.8<br>(17.7) | 36.6 (11.8) | 36.9 (12.6) | 36.3<br>(11.5) | 18.3<br>(18.6) | 18.4 (19.4) | 18.1 (19.4) |
| Number Age 9-13 | 1849<br>(1398) | 1689<br>(1423) | 2020<br>(1400) | 2356 (1297) | 2113 (1516) | 2628<br>(1027) | 1235<br>(1303) | 1144 (1173) | 1325<br>(1512) |
| Sum of Each Provider's FTE | 6.45<br>(4.49) | 5.40<br>(3.15) | 7.58<br>(5.48) | 8.33 (4.46) | 6.42 (3.50) | 10.47<br>(4.64) | 4.17<br>(3.46) | 4.08 (2.21) | 4.27 (4.58) |
| Medicaid Share (%) | 22.3<br>(27.1) | 21.5<br>(26.9) | 23.1<br>(28.3) | 21.7 (20.3) | 22.4 (19.7) | 20.9<br>(22.4) | 23.0<br>(34.4) | 20.3 (35.8) | 25.7 (35.6) |
| Participating Child Characteristics | (N=62246) | (N=28407) | (N=33839) | (N=40591) | (N=18447) | (N=22144) | (N=21655) | (N=9960) | (N=11695) |
| Age at Baseline | 8.10<br>(1.39) | 8.11<br>(1.39) | 8.08<br>(1.39) | 8.16 (1.38) | 8.18 (1.38) | 8.14<br>(1.39) | 7.98<br>(1.39) | 7.97 (1.39) | 7.99 (1.39) |
| Sex, n (%) |  |  |  |  |  |  |  |  |  |
| Female | 30183<br>(48.49%) | 13762<br>(48.45%) | 16421<br>(48.53%) | 19756<br>(48.67%) | 8939<br>(48.46%) | 10817<br>(48.85%) | 10427<br>(48.15%) | 4823<br>(48.42%) | 5604<br>(47.92%) |
| Male | 31869<br>(51.20%) | 14453<br>(50.88%) | 17416<br>(51.47%) | 20643<br>(50.86%) | 9318<br>(50.51%) | 11325<br>(51.14%) | 11226<br>(51.84%) | 5135<br>(51.56%) | 6091<br>(52.08%) |
| Unknown | 194<br>(0.31%) | 192<br>(0.68%) | 2 (0.01%) | 192 (0.47%) | 190 (1.03%) | 2 (0.01%) | 2 (0.01%) | 2 (0.02%) | 0 |

### Median age at vaccine completion (primary outcome) and vaccine initiation (**Table 2**)

**Table 2:** Kaplan-Meier Estimates for Median Age of Completion and Initiation and Completion and Initiation by 13 years.

| Sample | Outcome | Median Age |  |  | Cumulative by 13y |  |  |
| --- | --- | --- | --- | --- | --- | --- | --- |
|  |  | 9-10 | 11-12 | Difference (95% CI) | 9-10 | 11-12 | RR (95% CI) |
| Combined | Completion | 12.2 | 13.3 | -1.1 (-1.9, -0.3) | 61.7% | 45.1% | 1.37 (1.04, 1.74) |
|  | Initiation | 11.0 | 11.8 | -0.8 (-1.3, -0.1) | 77.2% | 67.0% | 1.15 (1.00, 1.32) |
| Colorado | Completion | 12.1 | 12.8 | -0.7 (-1.0, -0.2) | 66.7% | 54.5% | 1.22 (1.04, 1.45) |
|  | Initiation | 10.7 | 11.4 | -0.7 (-1.1, -0.2) | 81.1% | 74.2% | 1.09 (0.99, 1.21) |
| California | Completion | 13.0 | N/A | N/A | 50.9% | 25.3% | 2.01 (1.68, 2.77) |
|  | Initiation | 11.3 | 12.8 | -1.5 (-2.3, -0.9) | 69.3% | 52.6% | 1.32 (1.17, 1.64) |
| *Confidence intervals obtained using the nonparametric bootstrap, with resampling at the practice level stratified by study arm. |  |  |  |  |  |  |  |

The median age at series completion [primary outcome] was 12.2 years in the 9-10 group vs 13.3 years in the 11-12 group [difference of -1.1 years (95% confidence interval [CI], -2.0 to -0.3)]. The median age at initiation was 11.0 years in the 9-10 group vs 11.8 in the 11-12 group [difference of -0.8 years (95% CI, -1.3 to -0.2)].

### Kaplan-Meier curves **(Figure 2**)

**Figure 2:**
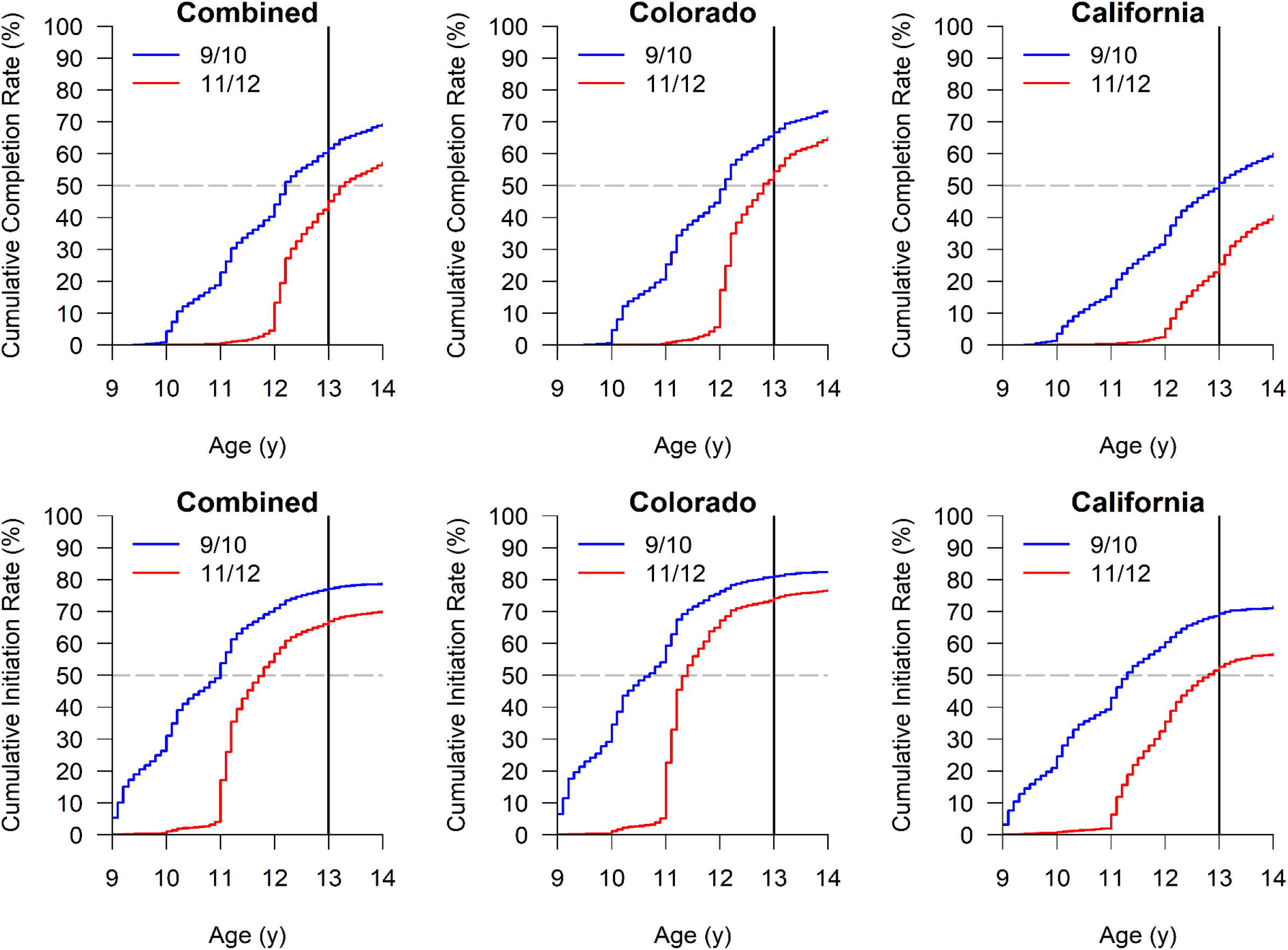
HPV Initiation and Completion Rates: Combined, Colorado and California.

The 9-10 intervention group had younger age at series completion [primary outcome] and initiation throughout the 4-year period. Differences between groups were greatest initially but persisted even by age 13. Results were similar between Colorado and California sites, and for each of the two California systems independently (**Figure S2**).

### Vaccination rates by age 13 years (**Table 2 and Figure 2**)

Completion rates by age 13 were 61.7% in the 9-10 group vs 45.1% in the 11-12 group [risk ratio (RR) 1.37 (95% CI, 1.04 to 1.74)]; initiation rates were 77.2% vs 67.0% [RR 1.15 (95% CI, 1.00 to 1.32)].

### Covariate-adjusted Cox Proportional Hazards Models (**Table 3**)

**Table 3.** Covariate-Adjusted Cox Proportional Hazards Models.

| **Outcome** |  | **HR (95% CI)** | **P** |
| --- | --- | --- | --- |
| Completion | Arm (9-10 v. 11-12) | 2.37 (1.94, 2.89) | **<0.001** |
|  | Arm x State (California v. Colorado) | 1.42 (0.83, 2.40) | 0.198 |
|  | Arm x Up-to-Date Rate (+10%) | 0.99 (0.81, 1.21) | 0.933 |
|  | Arm x Number Age 9-13 (+1000) | 1.10 (0.97, 1.23) | 0.129 |
|  | Arm x Medicaid Share (+10%) | 0.97 (0.91, 1.04) | 0.353 |
|  | Arm x Age (+1y) | 0.45 (0.40, 0.49) | **<0.001** |
|  | Arm x Sex (Female v. Male) | 1.08 (1.01, 1.16) | **0.036** |
| Initiation | Arm (9-10 v. 11-12) | 2.19 (1.84, 2.62) | **<0.001** |
|  | Arm x State (California v. Colorado) | 1.24 (0.76, 2.01) | 0.383 |
|  | Arm x Up-to-Date Rate (+10%) | 0.98 (0.81, 1.18) | 0.825 |
|  | Arm x Number Age 9-13 (+1000) | 1.08 (0.98, 1.20) | 0.130 |
|  | Arm x Medicaid Share (+10%) | 0.97 (0.91, 1.03) | 0.341 |
|  | Arm x Age (+1y) | 0.59 (0.54, 0.64) | **<0.001** |
|  | Arm x Sex (Female v. Male) | 1.04 (0.99, 1.10) | 0.114 |

\*Each row reports on a separate regression analysis. Models adjusted for state, up-to-date rate, number age 9-13, Medicaid share, age and sex, and inferences were clustered at the practice level.

This model estimated an adjusted Hazards Ratio (HR) of 2.37 (p<.001) for age at completion and 2.19 (p<.001) for age at initiation for the 9-10 versus 11-12 groups. Assessment of effect modification by prespecified subgroups showed no significant differences between state overall or by percentage of patients with Medicaid insurance within practices. There was slightly higher intervention effectiveness among female vs male adolescents for series completion (p=0.037), but not for initiation. The intervention was more effective for children who were younger [HR 0.44 (0.40 to 0.49) for each additional year] for completion and initiation [HR 0.59 (0.54 to 0.64)]. The sensitivity analysis limited to the children age 9-10 at baseline (**Figure S3**) demonstrated effects qualitatively consistent but quantitatively more modest than the primary analysis.

### Effect of intervention on delivery of other vaccines **(Table S1)**

Likelihood of receipt of the MenACWY vaccine during the study or rate of receipt by age 13 did not differ between study arms, There was a higher rate of receipt of Tdap during the study in the 9-10 study arm [HR 1.47 (CI 1.13, 1.94)]; rates of receipt by 13 years were similar.

## Discussion

The enormous promise of the HPV vaccine in preventing cancer has not been fully realized due to suboptimal initiation and completion of the series among adolescent populations. Our results demonstrate that initiating routine recommendations for HPV vaccination at ages 9-10 rather than at 11-12 years resulted in a median series completion age 1.1 years earlier and series completion by age 13 years more than 15 percentage points higher. Initiation of vaccination by age 13 years improved by approximately 10 percentage points [HR 2.52 (1.93, 3.27)]. These results, in addition to previously published data from our trial showing that clinicians found the change to an earlier initiation age to be highly feasible and acceptable^38,39^, support consideration of a preferential recommendation for initiation of HPV vaccination at age 9-10 years.

Our data suggest that earlier initiation at 9-10 years was not simply shifting the age of initiation among those who would eventually be vaccinated before age 13 but was increasing the percentage of adolescent who received the vaccine before 13 years of age. The timing of HPV vaccination prior to any sexual activity is critical. Data demonstrate that the rate of sexual activity increases rapidly after age 13 years^40^, underlying the importance of vaccination by this age. Our data demonstrating greater success in achieving full vaccination and in initiating vaccination among adolescents age 13 years may be key to preventing infection among younger adolescents before sexual activity.

The mechanism for effectiveness of this simple intervention is likely to be multi-factorial. We had hypothesized that earlier initiation might “uncouple” discussions of the vaccine as cancer prevention from sexual activity. Indeed, qualitative data from clinicians in our trial indicated that they perceived conversations were shorter with less push-back from parents in the 9-10 arm versus 11-12 arm.^39^ However, clinician surveys noted no differences in discussing sexual activity with patients in the 9-10 versus 11–12-year groups.^38^ Nonetheless, clinician perception that the discussion was easier among the 9–10-year-old group is key because clinicians who anticipate a difficult discussion may be less likely to make a strong recommendation.^41,42^ Also, perhaps offloading one of the adolescent vaccines to 9-10 years and decreasing the number of vaccines given simultaneously at 11-12 may have helped overcome parental hesitancy for HPV vaccine (which is often the vaccine forgone when adolescents need multiple vaccinations).^12,43,44^ Finally, earlier initiation likely gave more opportunities for repeated discussions between clinicians and parents, perhaps resulting in higher vaccine acceptance.

The World Health Organization in 2022 included an option for single-dose vaccination in some groups based on data suggesting durability of antibody levels and protection after a single dose. Our data show a substantially higher rate of initiation by 13 years in practices using an age 9-10 introduction. Thus, even if the US routine recommendation changes to one dose, our data suggest this will be better accomplished by introduction at 9-10 years.

Limitations include the setting of only two states. Although we had variation in types of practices (private, public and systems), all practices were urban or suburban; therefore, our data may not be representative of rural areas. Also, we couldn’t account for patients who left participating practices, although the amount of turnover of adolescents would likely be the same in intervention and control practices. Strengths include our ability to study HPV 9-10 in isolation of other interventions, high fidelity to study arms, the prospective randomized study design, adherence to the same training and follow-up protocols between different study arms, and the similarity of findings in all sites.

Currently, the American Academy of Pediatrics (AAP) recommends starting “at between 9 and 12□years, at an age that the provider deems optimal for acceptance and completion of the vaccination series.” Many states are following AAP guidelines^45^ and recent data suggest most pediatricians and family medicine physicians are following the recommendations of their national professional organizations. Combined with our qualitative^39^ and survey studies^38^ of clinicians that noted feasibility and acceptability of HPV 9-10 initiation, as well as our data showing either no detrimental effect or a positive effect on delivery of other vaccines routinely recommended at 11-12 years, results of this trial suggest that professional organizations and states should consider a preferential recommendation for HPV vaccine initiation at 9-10 with catch-up at ages 11-12. The 11–12-year-old vaccine platform could continue to be anchored by Tdap and MenACWY, as well as Influenza vaccine when seasonally appropriate. Our data may also be highly relevant to other countries that currently routinely recommend initiation later than age 9 years,^46^ although the effect of cultural differences and different delivery systems will need to be examined. Overall, early initiation at 9-10 years, given its simplicity, acceptability, and effectiveness, may prove to be an important factor in raising HPV vaccination rates among children and adolescents.

## Data Availability

All data produced are available to those that request it at

## References

1. U.S. Centers for Disease Control and Prevention. Cancers Linked with HPV Each Year. U.S. Centers for Disease Control and Prevention. Accessed September 1, 2026. https://www.cdc.gov/cancer/hpv/cases.html

2. Markowitz LE, Unger ER. Human Papillomavirus Vaccination. N Engl J Med. May 11 2023;388(19):1790–1798. doi:10.1056/NEJMcp2108502

3. de Martel C, Plummer M, Vignat J, Franceschi S. Worldwide burden of cancer attributable to HPV by site, country and HPV type. Int J Cancer. Aug 15 2017;141(4):664–670. doi:10.1002/ijc.30716

4. Schiller JT, Castellsagué X, Garland SM. A review of clinical trials of human papillomavirus prophylactic vaccines. Vaccine. Nov 20 2012;30 Suppl 5(5):F123–138. doi:10.1016/j.vaccine.2012.04.108

5. Paavonen J, Naud P, Salmerón J, et al. Efficacy of human papillomavirus (HPV)-16/18 AS04-adjuvanted vaccine against cervical infection and precancer caused by oncogenic HPV types (PATRICIA): final analysis of a double-blind, randomised study in young women. Lancet. Jul 25 2009;374(9686):301–314. doi:10.1016/s0140-6736(09)61248-4

6. Garland SM, Hernandez-Avila M, Wheeler CM, et al. Quadrivalent vaccine against human papillomavirus to prevent anogenital diseases. N Engl J Med. May 10 2007;356(19):1928–43. doi:10.1056/NEJMoa061760

7. Tsentemeidou A, Fyrmpas G, Stavrakas M, et al. Human Papillomavirus Vaccine to End Oropharyngeal Cancer. A Systematic Review and Meta-Analysis. Sex Transm Dis. Sep 1 2021;48(9):700–707. doi:10.1097/olq.0000000000001405

8. Rosado C, FernandesÂ R, Rodrigues AG, Lisboa C. Impact of Human Papillomavirus Vaccination on Male Disease: A Systematic Review. Vaccines (Basel). Jun 9 2023;11(6). doi:10.3390/vaccines11061083

9. Bhatla N, Muwonge R, Malvi SG, et al. Impact of age at vaccination and cervical HPV infection status on binding and neutralizing antibody titers at 10 years after receiving single or higher doses of quadrivalent HPV vaccine. Hum Vaccin Immunother. Dec 15 2023;19(3):2289242. doi:10.1080/21645515.2023.2289242

10. National Committee for Quality Assurance. HEDIS Measures and Technical Resources. National Committee for Quality Assurance. Accessed September 1, 2026. https://www.ncqa.org/hedis/measures/

11. Meites E, Kempe A, Markowitz LE. Use of a 2-Dose Schedule for Human Papillomavirus Vaccination -Updated Recommendations of the Advisory Committee on Immunization Practices. MMWR Morb Mortal Wkly Rep. Dec 16 2016;65(49):1405–1408. doi:10.15585/mmwr.mm6549a5

12. Pingali C, Yankey D, Elam-Evans LD, et al. Vaccination Coverage Among Adolescents Aged 13-17 Years -National Immunization Survey-Teen, United States, 2024. MMWR Morb Mortal Wkly Rep. Aug 14 2025;74(30):466–472. doi:10.15585/mmwr.mm7430a1

13. Cataldi JR, Suresh K, Brewer SE, et al. Boot Camp Translation using Community-Engaged messaging for adolescent Vaccination: A Cluster-Randomized trial. Vaccine. Feb 15 2024;42(5):1078–1086. doi:10.1016/j.vaccine.2024.01.042

14. Gilkey MB, Grabert BK, Heisler-MacKinnon J, et al. Coaching and Communication Training for HPV Vaccination: A Cluster Randomized Trial. Pediatrics. Aug 1 2022;150(2). doi:10.1542/peds.2021-052351

15. Szilagyi PG, Humiston SG, Stephens-Shields AJ, et al. Effect of Training Pediatric Clinicians in Human Papillomavirus Communication Strategies on Human Papillomavirus Vaccination Rates: A Cluster Randomized Clinical Trial. JAMA Pediatr. Sep 1 2021;175(9):901–910. doi:10.1001/jamapediatrics.2021.0766

16. Stephens AB, Wynn CS, Stockwell MS. Understanding the use of digital technology to promote human papillomavirus vaccination -A RE-AIM framework approach. Hum Vaccin Immunother. 2019;15(7-8):1549–1561. doi:10.1080/21645515.2019.1611158

17. Dempsey AF, Pyrznawoski J, Lockhart S, et al. Effect of a Health Care Professional Communication Training Intervention on Adolescent Human Papillomavirus Vaccination: A Cluster Randomized Clinical Trial. JAMA Pediatr. May 7 2018;172(5):e180016. doi:10.1001/jamapediatrics.2018.0016

18. Rand CM, Schaffer SJ, Dhepyasuwan N, et al. Provider Communication, Prompts, and Feedback to Improve HPV Vaccination Rates in Resident Clinics. Pediatrics. Apr 2018;141(4). doi:10.1542/peds.2017-0498

19. Rand CM, Tyrrell H, Wallace-Brodeur R, et al. A Learning Collaborative Model to Improve Human Papillomavirus Vaccination Rates in Primary Care. Acad Pediatr. Mar 2018;18(2s):S46–s52. doi:10.1016/j.acap.2018.01.003

20. Reno JE, O’Leary S, Garrett K, et al. Improving Provider Communication about HPV Vaccines for Vaccine-Hesitant Parents Through the Use of Motivational Interviewing. J Health Commun. 2018;23(4):313–320. doi:10.1080/10810730.2018.1442530

21. Brewer NT, Hall ME, Malo TL, Gilkey MB, Quinn B, Lathren C. Announcements Versus Conversations to Improve HPV Vaccination Coverage: A Randomized Trial. Pediatrics. Jan 2017;139(1). doi:10.1542/peds.2016-1764

22. O’Leary S, Pyrzanowski J, Lockhart S, et al. Impact of a Provider Communication Training Intervention on Adolescent Human Papillomavirus Vaccination: A Cluster Randomized, Clinical Trial. Open Forum Infectious Diseases. 10/01 2017;4:S61–S61. doi:10.1093/ofid/ofx162.144

23. Kempe A, O’Leary ST, Shoup JA, et al. Parental Choice of Recall Method for HPV Vaccination: A Pragmatic Trial. Pediatrics. Mar 2016;137(3):e20152857. doi:10.1542/peds.2015-2857

24. O’Leary ST, Lee M, Lockhart S, et al. Effectiveness and Cost of Bidirectional Text Messaging for Adolescent Vaccines and Well Care. Pediatrics. Nov 2015;136(5):e1220–7. doi:10.1542/peds.2015-1089

25. Kharbanda EO, Stockwell MS, Fox HW, Andres R, Lara M, Rickert VI. Text message reminders to promote human papillomavirus vaccination. Vaccine. 2011;29(14):2537–2541. doi:10.1016/j.vaccine.2011.01.065

26. Brewer SK, Stefanos R, Murthy NC, Asif AF, Stokley S, Markowitz LE. Human papillomavirus vaccination at age 9 or 10 years to increase coverage -a narrative review of the literature, United States 2014-2024. Hum Vaccin Immunother. Dec 2025;21(1):2480870. doi:10.1080/21645515.2025.2480870

27. Perkins RB, Humiston S, Oliver K. Evidence supporting the initiation of HPV vaccination starting at age 9: Collection overview. Hum Vaccin Immunother. 2023. vol. 3.

28. O’Leary ST. Why the American Academy of Pediatrics recommends initiating HPV vaccine at age 9. Hum Vaccin Immunother. Nov 30 2022;18(6):2146434. doi:10.1080/21645515.2022.2146434

29. Bednarczyk RA, Brandt HM. Descriptive epidemiology of age at HPV vaccination: Analysis using the 2020 NIS-Teen. Hum Vaccin Immunother. Dec 31 2023;19(1):2204784. doi:10.1080/21645515.2023.2204784

30. Kong WY, Huang Q, Thompson P, Grabert BK, Brewer NT, Gilkey MB. Recommending Human Papillomavirus Vaccination at Age 9: A National Survey of Primary Care Professionals. Acad Pediatr. May-Jun 2022;22(4):573–580. doi:10.1016/j.acap.2022.01.008

31. Holder N, Smith K, Coyne-Beasley T. The Adolescent Immunization Platform: The Past and Future. J Adolesc Health. Aug 2025;77(2S):S11–S13. doi:10.1016/j.jadohealth.2025.05.002

32. Immunization of adolescents. Recommendations of the Advisory Committee on Immunization Practices, the American Academy of Pediatrics, the American Academy of Family Physicians, and the American Medical Association. MMWR Recomm Rep. Nov 22 1996;45(RR-13):1–16.

33. Bilukha OO, Rosenstein N, National Center for Infectious Diseases, Centers for Disease Control and Prevention. Prevention and control of meningococcal disease. Recommendations of the Advisory Committee on Immunization Practices (ACIP). MMWR Recomm Rep. May 27 2005;54(RR-7):1–21.

34. Broder KR, Cortese MM, Iskander JK, et al. Preventing tetanus, diphtheria, and pertussis among adolescents: use of tetanus toxoid, reduced diphtheria toxoid and acellular pertussis vaccines recommendations of the Advisory Committee on Immunization Practices (ACIP). MMWR Recomm Rep. Mar 24 2006;55(RR-3):1–34.

35. Markowitz LE, Dunne EF, Saraiya M, et al. Quadrivalent Human Papillomavirus Vaccine: Recommendations of the Advisory Committee on Immunization Practices (ACIP). MMWR Recomm Rep. Mar 23 2007;56(RR-2):1–24.

36. Child and Adolescent Health Measurement Initiative. 2003-2004 National Survey of Children’s Health (NSCH) data query. Data Resource Center for Child and Adolescent Health, supported by Cooperative Agreement from the U.S. Department of Health and Human Services, Health Resources and Services Administration (HRSA), Maternal and Child Health Bureau (MCHB). Accessed September 19, 2026. http://childhealthdata.org

37. Brewer SE, Crump NM, O’Leary ST. Patient-Centered Research Priorities: A Mixed-Methods Approach from the Colorado Children’s Outcomes Network (COCONet). J Am Board Fam Med. 2019;32(5):674–684. doi:10.3122/jabfm.2019.05.190028

38. Szilagyi PG, Gurfinkel D, Clark E, et al. Clinician Perceptions of the Impact of Switching HPV Vaccine Initiation to 9-10 Years. Pediatrics. May 1 2026;157(5). doi:10.1542/peds.2025-074674

39. Tietbohl CK, Gurfinkel D, Duran D, et al. Feasibility and Acceptability of Recommending HPV Vaccine at Ages 9–10 Years. Pediatrics. Jul 1 2025;156(1). doi:10.1542/peds.2024-069625

40. Guttmacher Institute. Adolescent Sexual and Reproductive Health in the United States. Guttmacher Institute. Accessed December 1, 2025, https://www.guttmacher.org/sites/default/files/factsheet/adolescent-sexual-and-reproductive-health-in-united-states.pdf

41. Lin C, Mullen J, Smith D, Kotarba M, Kaplan SJ, Tu P. Healthcare Providers’ Vaccine Perceptions, Hesitancy, and Recommendation to Patients: A Systematic Review. Vaccines (Basel). Jul 1 2021;9(7). doi:10.3390/vaccines9070713

42. Gilkey MB, Malo TL, Shah PD, Hall ME, Brewer NT. Quality of physician communication about human papillomavirus vaccine: findings from a national survey. Cancer Epidemiol Biomarkers Prev. Nov 2015;24(11):1673–9. doi:10.1158/1055-9965.epi-15-0326

43. Ryan G, Ashida S, Gilbert PA, et al. The Use of Medical Claims Data for Identifying Missed Opportunities for HPV Immunization Among Privately Insured Adolescents in the State of Iowa. J Community Health. Oct 2022;47(5):783–789. doi:10.1007/s10900-022-01110-7

44. Espinosa CM, Marshall GS, Woods CR, et al. Missed Opportunities for Human Papillomavirus Vaccine Initiation in an Insured Adolescent Female Population. J Pediatric Infect Dis Soc. Nov 24 2017;6(4):360–365. doi:10.1093/jpids/pix067

45. Kates J, Bell C. State Recommendations for Routine Childhood Vaccines: Increasing Departure from Federal Guidelines. KFF. Accessed May 27, 2026, https://www.kff.org/state-health-policy-data/state-recommendations-for-routine-childhood-vaccines-increasing-departure-from-federal-guidelines/

46. World Health Organization. Vaccination schedule for Human Papillomavirus (HPV). World Health Organization. Accessed September 1, 2026. https://immunizationdata.who.int/global/wiise-detail-page/vaccination-schedule-for-human-papilloma-virus?ISO_3_CODE=&TARGETPOP_GENERAL=

