## Supplemental Figures and Table for "Effect of HPV Vaccine Introduction at 9-10 years: A Randomized Clinical Trial"

**Supplemental Materials**


| Figure Number | Title | Page |
|---|---|---|
| Figure S1 | Electronic Health Record (EHR) captured opportunities (HPV doses) for 9-10 yos by 6-month intervals, among Well Child Care visits by state | 2 |
| Figure S2 | HPV Vaccine Completion and Initiation Rates by Health System in California--Children's Hospital of Orange County (CHOC) and UCLA Health Clinics (UCLA) | 3 |
| Figure S3 | HPV Vaccine Completion and Initiation Rates of Baseline Cohort and Cox Models | 4 |
| Table S1 | Receipt During Study and Up-to-date by Age 13 Years for Other Vaccines Recommended at 11-12 Year Platform, by Trial Arm | 5 |

**Figure S1: EHR captured opportunities (HPV doses) for 9-10 yos by 6-month intervals, among Well Child Care visits by state**

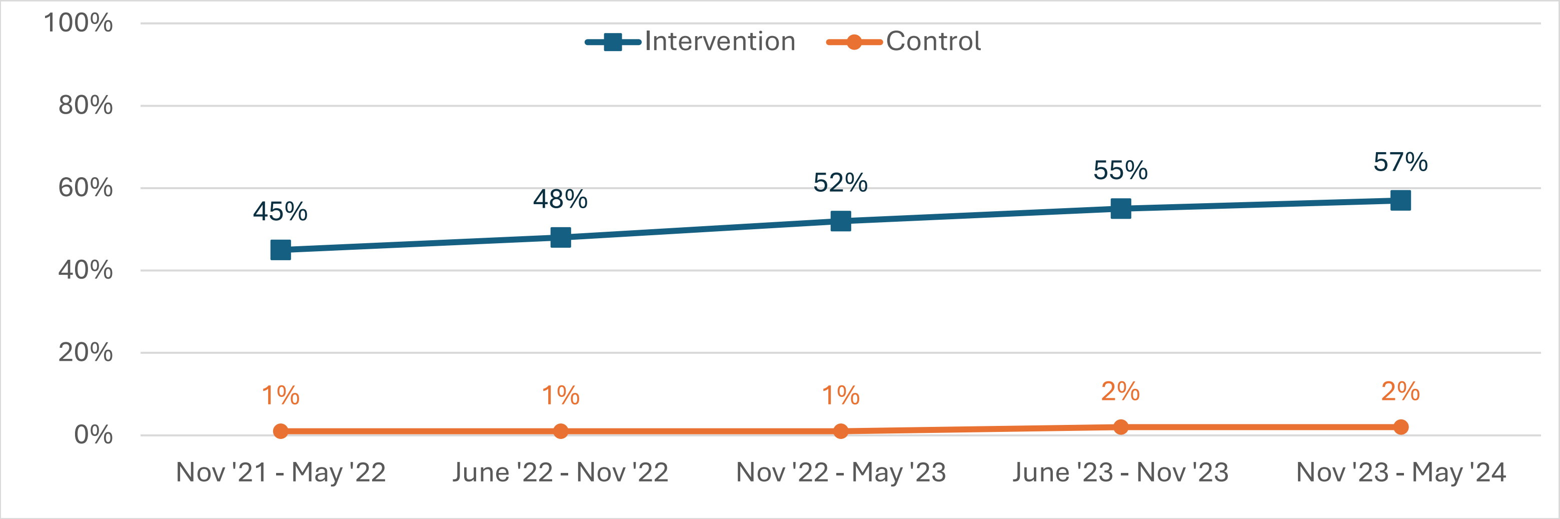


**Colorado practices**

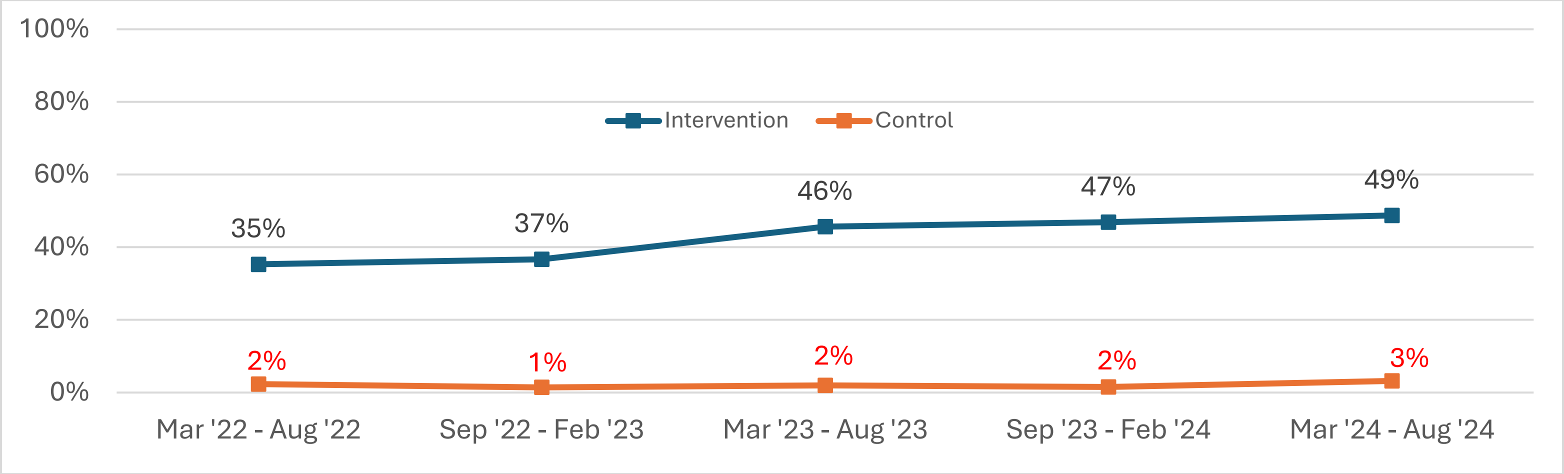


**California practices**

**Figure S2: HPV Vaccine Completion and Initiation Rates by Health System in California--Children's Hospital of Orange County (CHOC) and UCLA Health Clinics (UCLA)**

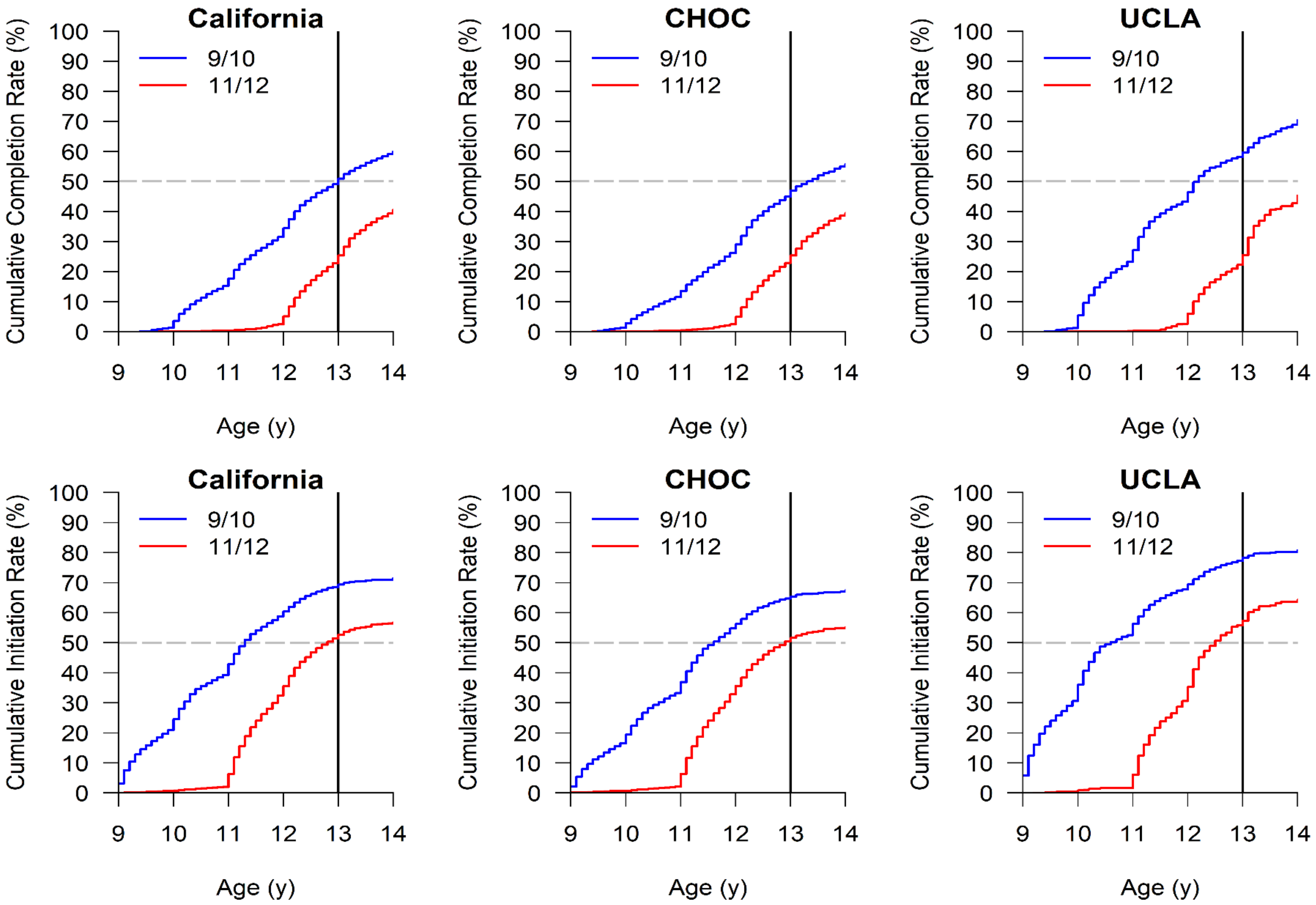

**Figure S3: HPV Vaccine Completion and Initiation Rates for Baseline Cohort Only**

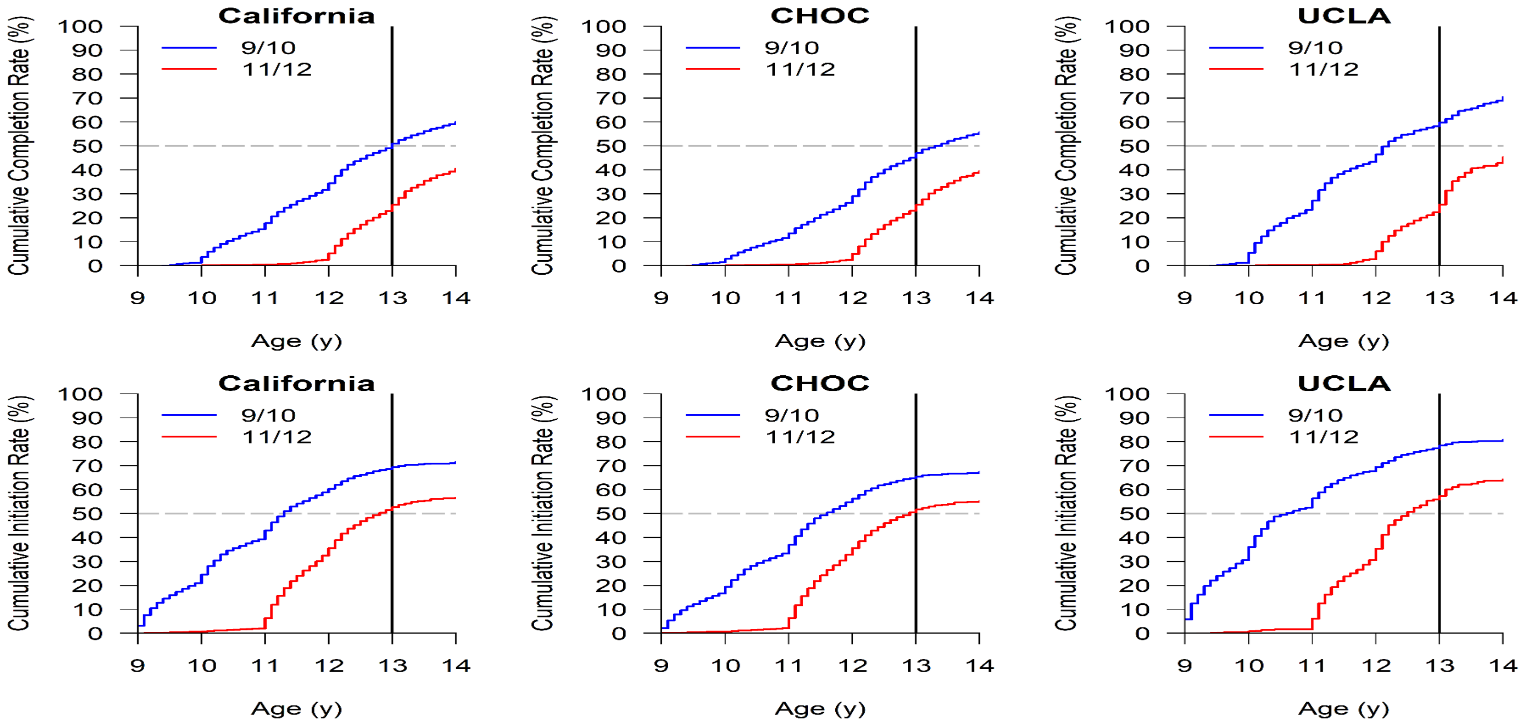


### Cox Models for Baseline Cohort

| Outcome | | HR (95% CI) | P |
|---|---|---|---|
| Completion | Arm (9-10 v. 11-12) | 1.39 (1.15, 1.67) | **<0.001** |
| Initiation | Arm (9-10 v. 11-12) | 1.40 (1.19, 1.64) | **<0.001** |
| *Each row reports on a separate regression analysis. Models adjusted for state, up-to-date rate, number age 9-13, Medicaid share, age and sex, and inferences were clustered at the practice level. | | | |

**Table S1: Receipt During Study and Up to Date by Age 13 Years for Other Vaccines Recommended at 11-12 Year Platform, by Trial Arm**

| | Receipt of Other Vaccinations | | Up-to-Date Status at Age 13.0 Years | | |
|---|---|---|---|---|---|
| **Vaccination** | **Hazard Ratio (95% CI)[a]** | **P Value[a]** | **HPV 9-10 Arm, %** | **HPV 11-12 Arm, %** | **Risk Ratio (95% CI)[b]** |
| MenACWY | 1.05 (0.77, 1.44) | 0.74 | 76.0 | 71.0 | 1.07 (0.82, 1.34) |
| Tdap | 1.47 (1.13, 1.92) | 0.004 | 86.1 | 83.8 | 1.03 (0.94, 1.13) |

*Abbreviations:* CI, confidence interval; HPV, human papillomavirus; MenACWY, Meningococcal; Tdap, tetanus-diphtheria-acellular pertussis.

[a] Hazard ratio and P value from a Cox proportional hazards regression model for time to receipt of the indicated vaccination, comparing the HPV 9-10 arm with the HPV 11-12 arm; the model was adjusted for patient age, sex, state, and practice-level randomization balancing factors, consistent with the primary analysis.

[b] Risk ratio for the proportion of children up to date on the indicated vaccination at age 13.0 years, comparing the HPV 9-10 arm with the HPV 11-12 arm; 95% CIs were derived using a cluster bootstrap algorithm to account for clustering by practice.
